# *XPOT* Deficiency causes a human disorder through impaired tRNA nuclear export

**DOI:** 10.64898/2026.01.28.26344748

**Authors:** S. von Hardenberg, I. Niehaus, A. Wiemers, T. Rothoeft, K. Huang, C. Petree, V. Schäffer, C. Phillipe, A-L. Bruel, K. Warnatz, M. Zamani, R. Ahmadi, A. Sedaghat, S. Bahram, S. Sedighzadeh, S. Ebrahimi, S. Khalilian, S. Landwehr-Kenzel, N. Schwerk, A. Elsayed, J. Rösler, S-J. Lin, S. Sabu, N. Strenzke, G. Sogkas, B. Vona, G. K. Varshney, N. Di Donato, B. Auber

## Abstract

**Background:** The transport of transfer RNAs (tRNAs) from the nucleus to the cytoplasm is a crucial step in the regulation of gene expression and protein synthesis. This process is mediated by specialized export molecules, among which XPOT (Exportin-t, XPO3) plays a central role by recognizing and transporting mature tRNAs through the nuclear pore complex. XPOT is not essential in RNA trafficking in the simple organisms, however the potential impact of XPOT deficiency in human health remains unresolved.

**Methods:** We identified eight patients from five unrelated families with rare biallelic germline variants in XPOT resulting in putative loss-of-function. Functional analyses were carried out in patient-derived fibroblasts, lymphoblastoid cells and zebrafish models. *Ex vivo* immunohistochemical stainings for Xpot were performed in the mouse cochlea. *xpot* knockout zebrafish models were generated to assess the morphology and hearing ability.

**Results:** All patients presented with a uniform clinical phenotype that included increased susceptibility to infection, bronchiectasis, severe sensorineural hearing loss, developmental delay, and growth retardation. We demonstrated a complete absence of *XPOT* protein expression in three patient-derived cell lines. *XPOT* deficiency leads to disruptions in protein synthesis of the cytokine TNFα pathway upon cellular stimulation. Additional XPO1 inhibition in XPOT deficient cells had little effect on cellular functions, suggesting alternative tRNA nuclear transporter pathways. Increased XPOT immunoreactivity was observed in type I spiral ganglion neurons and hair cells of the mouse cochlea, with enrichment in stereocilia. *xpot* knockout zebrafish model showed dysmorphic features, and reduced hearing, recapitulating key patient phenotypes.

**Conclusions:** Our findings establish a direct connection between impaired XPOT-dependent tRNA export and human pathology. It illustrates that perturbations in nuclear export pathways lead to disease. It also raises the possibility that other nuclear transport receptors may play similarly underappreciated roles in human health and disease. The identification of *XPOT* as a disease-associated gene opens up new research directions and potential targets for therapeutic intervention.

## Introduction

Transfer ribonucleic acids (tRNAs) are essential components of the translational machinery, delivering amino acids to ribosomes and enabling the accurate decoding of messenger RNA (mRNA) into polypeptide chains^1^. In eukaryotes, tRNAs are transcribed in the nucleus as precursor molecules (pre-tRNAs) and undergo a series of maturation events, including removal of 5’ and 3’ extensions, addition of the 3’-terminal CCA sequence, covalent nucleoside modifications, and – in many cases – endonucleolytic splicing. Only fully processed tRNAs are exported from the nucleus, a quality-control checkpoint that ensures translational fidelity^2^.

Beyond this pathway, tRNAs also undergo bidirectional nucleocytoplasmic shuttling, which contributes to several layers of gene expression regulation^3, 4, 5^. As tRNAs are smaller than the 40 kD threshold for passive diffusion across the nuclear pore complex (NPC), their export from the nucleus is an active process^6^. The principal nuclear export receptor in eukaryotic cells is the conserved β-importin family member XPOT (Exportin-t, XPO3), which selectively binds fully matured tRNA^7, 8^ and transports it across the NPC following a nuclear export signal (NES). This process is regulated by the small GTPase Ran, which cycles between a nuclear GTP-bound and a cytoplasmic GDP-bound state^9^. The trimeric complex traverses the NPC and is disassembled in the cytoplasm following the hydrolysis of RanGTP.

tRNA subcellular trafficking is responsive to metabolic status, stress signals and other environmental cues.^10, 11^ Perturbations in tRNA processing, modification, or aminoacylation are increasingly recognized as causes of human neurodevelopmental and multisystem disorders, arising from pathogenic variants in mitochondrial tRNAs^12^, defects in tRNA processing or modification enzymes such as *CLP1*^13^, and in aminoacyl-tRNA synthetases (ARSs), underscoring their importance not only in translation, but also in maintaining cellular homeostasis.^2^ Additionally, many diseases without a primary genetic link to tRNA biology exhibit secondary alterations in tRNA abundance or maturation. Despite the central importance of nucleocytoplasmic tRNA transport, no Mendelian disease has previously been attributed to dysfunction of a tRNA nuclear transport.

Here, we delineate a previously undescribed autosomal recessive disorder linked to the tRNA exportin, *XPOT*. We present eight affected children from five unrelated families who share a uniform syndromic phenotype. Common manifestations include an increased susceptibility to infection, non-cystic fibrosis bronchiectasis, sensorineural hearing loss, global developmental and motor delays and growth retardation. Further analyses revealed that the identified variants lead to the absence of XPOT expression and most likely result in impaired nuclear export of mature tRNAs, significantly altering protein translation efficiency.

Taken together, these findings establish biallelic *XPOT* variants as the basis of a novel tRNA export-related disorder with a robust gene–phenotype relationship, highlighting the essential role of tRNA export for human health and expanding the landscape of tRNA-related disorders beyond previously recognized defects in tRNA modification and aminoacylation.

## Results

Family 1 consists of three affected children (P1, P2, P3) born to first-degree cousins (Figure 1A, Family 1). All three affected individuals presented with hearing impairment, recurrent (respiratory) infections, non-cystic fibrosis bronchiectasis, global developmental and motor delays and growth retardation. WGS revealed a homozygous germline variant, NM_007235.6(XPOT):c.2409_2412del p.(Thr804Serfs*6) in P1, P2 and P3 that was heterozygous in unaffected parents (F1-I:1 and F1-I:2) and not present in a healthy sibling (F1-II:2) suggesting an autosomal recessive mode of inheritance. No other plausible variants were identified, particularly not in the *CFTR* gene or in any gene known to cause primary ciliary dyskinesia.

**Figure 1:**
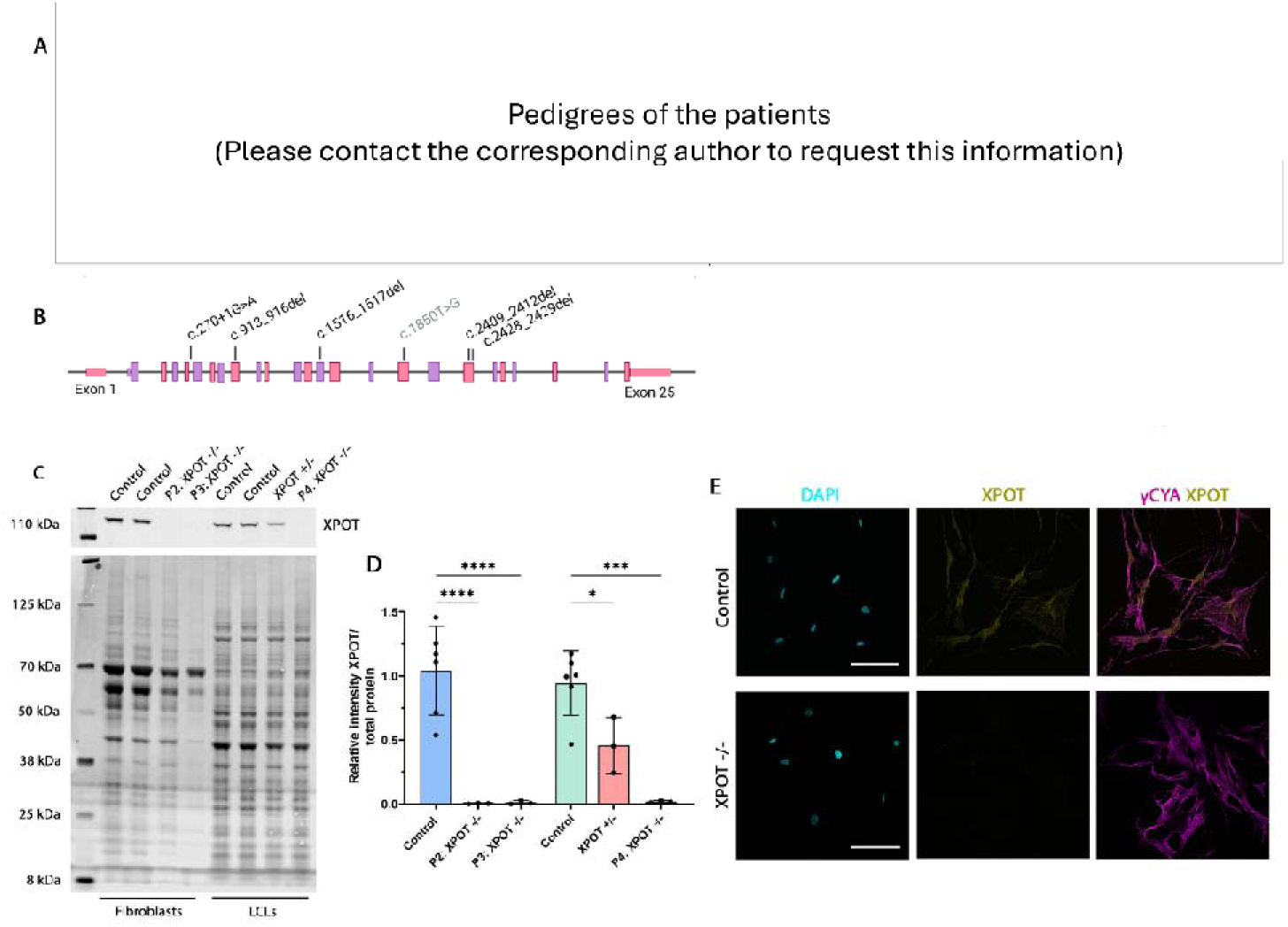
A: Pedigree of five families segregating autosomal recessive *XPOT* deficiency. Double lines indicate a consanguineous marriage. Filled black symbols indicate affected individuals, grey symbols indicate family members who were not tested, white symbols indicate individuals with two normal alleles, and half-shaded symbols represent heterozygous carriers. **B**: Schematic representation of the human *XPOT* gene structure comprising 25 exons, 24 of which are protein-coding (exon 2-25), including all variants identified here, as well as the missense variant c.1850T>G that has been observed in patients with a similar phenotype (grey)^16^. **C**: Representative western blot showing XPOT expression in control and patients’ fibroblasts (P2: XPOT-/- and P3: XPOT-/-) and LCLs (XPOT+/- and P4: XPOT-/-). Protein loading was verified by total protein staining. **D**: Quantification of XPOT expression in control and patients’ fibroblasts (P2 and P3) and LCLs (XPOT+/-and P4) normalized to total protein. Data are the mean of three independent experiments. The expression data of the control samples were combined, respectively. Error bars indicate SD; *, P < 0.05; ***, P < 0.001; ****, P < 0.0001 (one-way ANOVA). **E**: Double immunofluorescence for XPOT (yellow) and CYA (magenta) combined with DAPI staining (cyan) of control and patients’ fibroblasts (P2). Scale bars, 100 μm

Subsequently, five additional affected individuals from four unrelated families were identified (Figure 1A, Family 2, 3, 4, 5). In total, the cohort comprises eight individuals (seven males and one female) from five unrelated families, all exhibiting a remarkably uniform syndrome.

We identified four different homozygous frameshift variants (c.913_916del, c.2409_2412del, c.2428_2429del and c.1516del) resulting in premature stop-codon and presumable nonsense mediated decay and one homozygous canonical splice site variant NC_000012.12(NM_007235.6):c.270+1G>A in the *XPOT* gene (Figure 1B and Supplement Table S3). All homozygous variants are absent or extremely rare in the population (gnomAD v4.1.0) (Supplement Table S3).

*XPOT* demonstrates a probability of loss of function intolerance (pLI) score of 1, indicating a high intolerance to loss-of-function variants. The splice-site variant is unanimously predicted to abolish the splice donor site (SpliceSiteFinder-like, MaxEntScan, NNSPLICE, GeneSplicer, SpliceAI Visual (AI= 0.99), AbSplice). RNA analysis demonstrated skipping of exon 5 that would cause a frameshift (r.203_272del p.(Tyr68Cysfs*2)) (Supplement Figure S1).

To investigate XPOT protein expression in patient derived cells (primary dermal fibroblasts (P2 and P3) and lymphoblastoid cell lines (LCLs) (P4)), we performed western blot analysis and immunofluorescence staining. Cells from four healthy individuals were used as controls (2x fibroblast and 2x LCLs). Furthermore, protein expression of one heterozygous carrier was analyzed (XPOT +/-). Mutant cells did not express XPOT protein (Figure 1C and 1D). These observations were supported by immunofluorescence staining of mutant fibroblasts compared to WT fibroblasts (Figure 1E).

### *XPOT* deficiency results in a distinct syndromic disorder

The cohort comprises eight individuals aged between 5 and 41 years. Birth and neonatal histories were largely unremarkable, although one patient exhibited postnatal cyanotic episodes. Short stature was common(7/8). Facial phenotype (available for seven patients; Figure 2A und 2B), demonstrated distinct and recognisable gestalt: elongated face (6/7), particularly for the slightly older individuals, midface hypoplasia and broad nasal bridge (5/7), elongated, elapsed and wide philtrum, thin upper lip with thick lower (7/7), enophthalmos (4/7), low-hanging columella (4/7), and laterally tapering curved eyebrows (6/7). Pairwise rank analysis performed using GestaltMatcher^14^ confirmed significant similarity (please contact the corresponding author to request access to these data).

**Figure 2:**
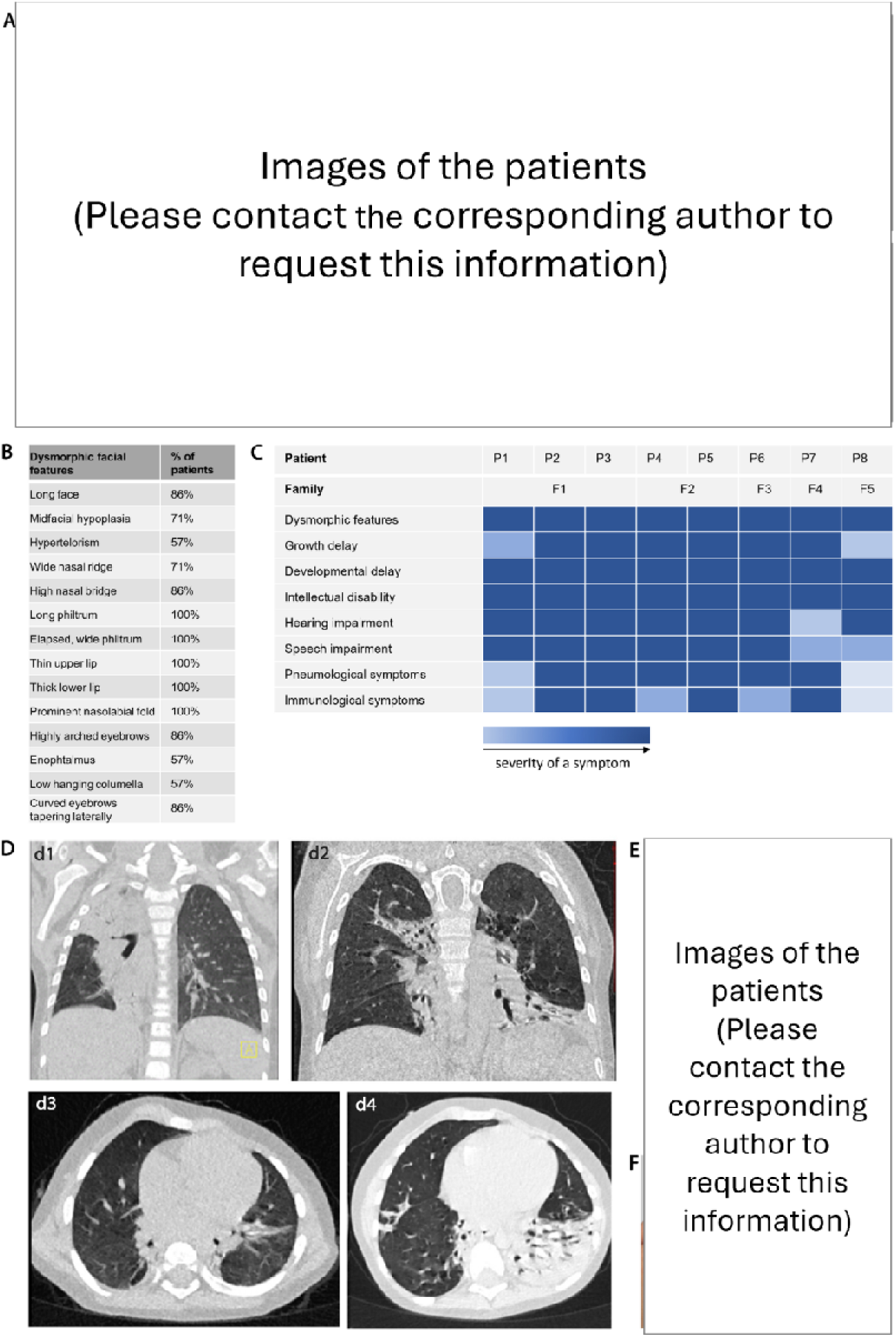
A: Facial images of the patients. Note the elongated face, midface hypoplasia, wide philtrum, thin upper lip, and thick lower lip. **B:** Overview of dysmorphic facial features observed in the study cohort. Percentages indicate the frequency of each feature within the cohort. **C:** Symptoms of individual patients displayed according to severity. Increasing color intensity corresponds to increased severity, (subjective assessments of the available clinical information). **D:** Representative sections from serial chest CT scans. Coronary CT sections of with subtotal consolidations of the right upper lobe and parts of the right lower lobe with positive bronchogram. The remaining lung parenchyma is unremarkable. In particular, there is no evidence of interstitial lung disease or fibrosis (**d1**). Coronary CT section of patient P2 with subtotal consolidations and bronchiectasis of the right upper lobe, the left lower lobe and parts of the right lower lobe (**d2**). Transversal CT sections of patient P2 (**d3**, **d4**) showing progression of consolidations and bronchiectasis throughout the left lower lobe. In all CT scans the remaining parenchyma is unremarkable. There is no evidence of interstitial lung disease or fibrosis. **E:** Psoriasiform lesions on the lower extremities of P2. **F:** Digital clubbing in P2, consistent with chronic tissue hypoxia.

Clinical findings are summarized in Figure 2C. A more detailed description of the patients is provided in supplemental information (see “Detailed description of audiological development in patients with XPOT deficiency”) or contact the corresponding author to request access to these data.

All patients exhibited developmental, speech and motor delay, with fine motor deficits and impaired coordination. Hearing impairment was reported in all cases and was typically severe or profound (with the exception of P7 who was only mildly or moderately affected). For P2 and P3, hearing loss was documented as mild/moderate at birth, but progressed to profound deafness over time. However, due to limited available information, normal hearing at birth could not be verified for all individuals. Computer tomography and/or MRI scans in P1, P2 and P3 revealed a normal auditory anatomy of the temporal bones and auditory nerves.

Detailed information regarding infections, skin diseases, and immunological alterations was available for 6 individuals and included recurrent viral or bacterial infections (6/6), psoriasis or eczema (4/6) and frequent oral ulcerations (5/6). Immunological workup revealed an elevated IgG/IgG4 in 6/6 cases. Vaccine titers and other immunoglobulins were within the normal range (6/6).

Pulmonary disease was severe and present in all individuals, including severe recurrent bronchitis and obstructive ventilation disorder. Available serial chest CT scans of five patients demonstrated consolidations, infiltrates, atelectasis, and bronchiectasis, while the remaining parenchyma was consistently unremarkable (Figure 2D). There was no evidence of interstitial lung disease or fibrosis, although ground-glass opacities with regional hyperinflation indicated possible regional hyperinflation due to small airways obstruction in some cases (Figure 2D). Ciliary diagnostics in (P2 and P3 revealed no evidence of ciliary dysfunction.

### XPOT is enriched in the mouse cochlea

Given that patients developed sensorineural hearing loss, we examined Xpot expression in the mouse cochlea. Visualization of published single-cell RNA-seq datasets in the gEAR portal showed that *Xpot* is expressed broadly across the embryonic (E14) and postnatal (P1, P7) mouse cochlear epithelium. The signal was present in sensory hair-cell populations and multiple supporting-cell classes at each stage, with no cluster lacking detectable transcripts. (Supplement Figure S2). Immunofluorescence stainings of mouse cochleae with two different anti-XPOT antibodies followed by confocal microscopy demonstrated a broad cytosolic and nuclear expression of XPOT. XPOT was particulary enriched in cells with high metabolic activity e.g. type 1 spiral ganglion neurons, hair cells, and in the stria vascularis. Moreover, strong XPOT staining was observed in the stereocilia of inner and outer hair cells, pointing to a potential additional role of XPOT in a specialized subcellular localization which is not involved in translation (Figure 3A-C, Supplement Figure S3A-C).

**Figure 3:**
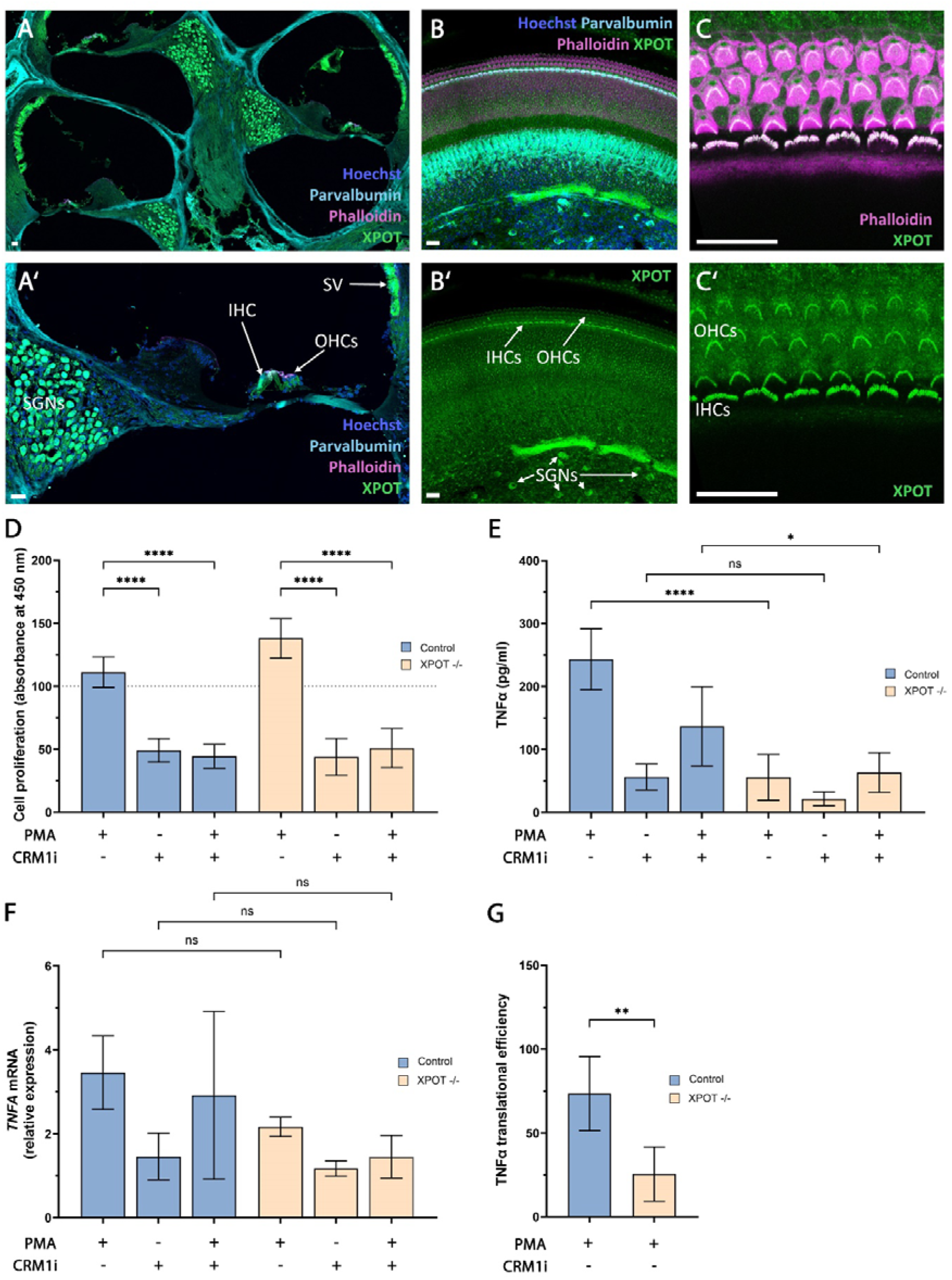
**A, B**: Immunostaining of a cryosection (A, p29) and a whole mount preparation (B, p5) of a wild type mouse cochlea (p29), demonstrating the expression of XPOT (green) in inner (IHC) and outer (OHC) hair cells, type I single ganglion neurons (SGNs) and stria vascularis (SV). **C:** High-magnification image from (B), focusing on the stereocilia of the one row of IHCs and the three rows of OHCs, strongly co-stained with anti-XPOT (green) and phalloidin (magenta). The kinocilium remained unstained. There were no obvious tonotopic or age-dependent differences in XPOT expression. In A and B, nuclei are stained with Hoechst (blue). Type I spiral ganglion neurons and inner hair cells are counterstained for parvalbumin (cyan) and actin filaments in stereocilia are stained with fluorophore-coupled phalloidin (magenta). All scale bars in A-C are 20 µm. **D:** Quantification of fibroblast proliferation after PMA, Selinexor or combined PMA and Selinexor treatment as a percentage relative to DMSO-treated controls. Data are the mean of four control cell lines and two patients cell lines (P2, P3), each from three independent assays. Error bars indicate SD; ****, P < 0.0001 (one-way ANOVA). **E**: Quantification of TNFα levels in fibroblasts by ELISA following treatment with PMA, Selinexor, or a combination of PMA and Selinexor. Data are the mean of four control cell lines and two patients cell lines (P2, P3), each from two independent assays. Error bars indicate SD; *, P < 0.05; ****, P < 0.0001, ns, not significant; (one-way ANOVA). **F**: qPCR analysis of TNFα mRNA expression normalized to GAPDH, after treatment with PMA, Selinexor, or PMA combined with Selinexor. Data are the mean of four control cell lines and two patients cell lines (P2, P3), each from two independent assays. Error bars indicate SD; ns, not significant (Kruskal-Wallis test). **G**: Relationship between *TNFA* mRNA and TNFα protein in patient-derived and control cells after PMA treatment. Data are the mean of four control cell lines and two patients cell lines (P2, P3), each from two independent assays. Error bars indicate SD; **, P < 0.05; (unpaired t-test).

### Significantly altered gene expression patterns in XPOT deficient fibroblasts

We performed transcriptome sequencing of patients derived dermal fibroblasts (P2 and P3) under steady state condition. First, we observed clear separation between *XPOT* deficient and control group in the cluster analysis (data not shown). Next, we identified 933 differentially expressed genes compared to control group (LogFoldChange <2 or >2, p-value adjusted <0.01, base expression of patients >100) (Supplement Figure S3D). As expected, we observed a marked downregulation of *XPOT* expression in the patients’ samples. Gene onthology enrichment analysis of differentially expressed genes showed a consistent overrepresentation of pathways linked to extracellular matrix organization and cellular stress/injury processes. (Supplement Figure S3E). Interestingly, alternative exporters potentially capable of compensating for *XPOT* function, e.g. XPO1 (CRM1) and XPO5 were not upregulated under steady-state conditions in patient-derived cells.

### CRM1 inhibition does not intensify translational dysfunction in patient cells

To assess the effect of XPOT on fibroblast proliferation and apoptosis, we performed proliferation and caspase assays with and without prior Phorbol 12-myristate 13-acetate (PMA) stimulation. No significant differences were observed between patient-derived cells and control fibroblasts (Figure 3D and Supplement Figure S3G). Since the nuclear export protein CRM1 (XPO1) transports a far greater variety of cargoes than any other exportin^15^, we investigated whether CRM1 could compensate for the absence of XPOT in patients’ cells. We subsequently evaluated proliferation and apoptosis after pretreatment with Selinexor, a selective inhibitor of CRM1, that binds to CRM1 and blocks its activity. In both groups, CRM1 inhibition (with or without prior PMA stimulation) significantly reduced cell proliferation, without significant effect on apoptosis. However, we saw no significant differences between patient-derived and control fibroblasts (Figure 3D and Supplement Figure S3G).

Given the patients’ increased susceptibility to infections, we compared the transcription and translation of the cytokine TNFα between patient-derived and control fibroblasts. By ELISA, we found that patient cells produced lower amounts of TNFα than control cells, both in the presence and absence of CRM1 inhibition. By contrast, only minor differences were detected at the mRNA level (Figure 3E and 3F). Figure 3G depicts the relationship between TNFA mRNA (x-axis, relative expression) and TNFα protein (y-axis, pg ml□¹) in patient-derived and control cells. Both groups exhibit a positive correlation; however, controls show a steeper regression (R² ≈ 0.864) than patient cells (R² ≈ 0.624). This flattened slope in patient cells reveals a clear deficit in protein produced per transcript unit, indicating substantially reduced translational output.

Collectively, these results demonstrate a significantly reduced translation efficiency in *XPOT*-deficient patient cells which is not further affected by inhibition of the CRM1 pathway.

To test the hypothesis that reduced protein expression is not attributable to diminished abundance of ARSs, we interrogated their transcript levels in the RNA-seq data. None of the ARSs were significantly decreased in patient cells compared to controls (Supplement Figure S3F).

### Zebrafish *xpot* knockout recapitulates key features of the human phenotype

The zebrafish ortholog of human *XPOT* shares significant sequence homology (89% similarity, 78% identity). We first characterized the spatiotemporal expression pattern of *xpot* during zebrafish development. Whole-mount *in situ* hybridization (WISH) revealed broad *xpot* expression throughout early development, with particularly prominent signal in the head region, including the otic vesicle at 1 day post fertilization (dpf) (Figure 4A, B). At 3 dpf, expression is restricted to the brain, eyes, and the developing inner ear (Figure 4C, D, arrowheads). By 5 dpf, *xpot* expression persisted in cranial structures, including the otic region (Figure 4E, F, arrowheads). Quantitative RT-PCR analysis confirmed robust *xpot* expression across developmental stages, with peak expression at 3 dpf (Figure 4G-i), a critical period of inner ear maturation. Tissue-specific analysis in adult zebrafish revealed *xpot* expression across multiple tissues, with the highest levels in the utricle (Figure 4G-ii).

**Figure 4:**
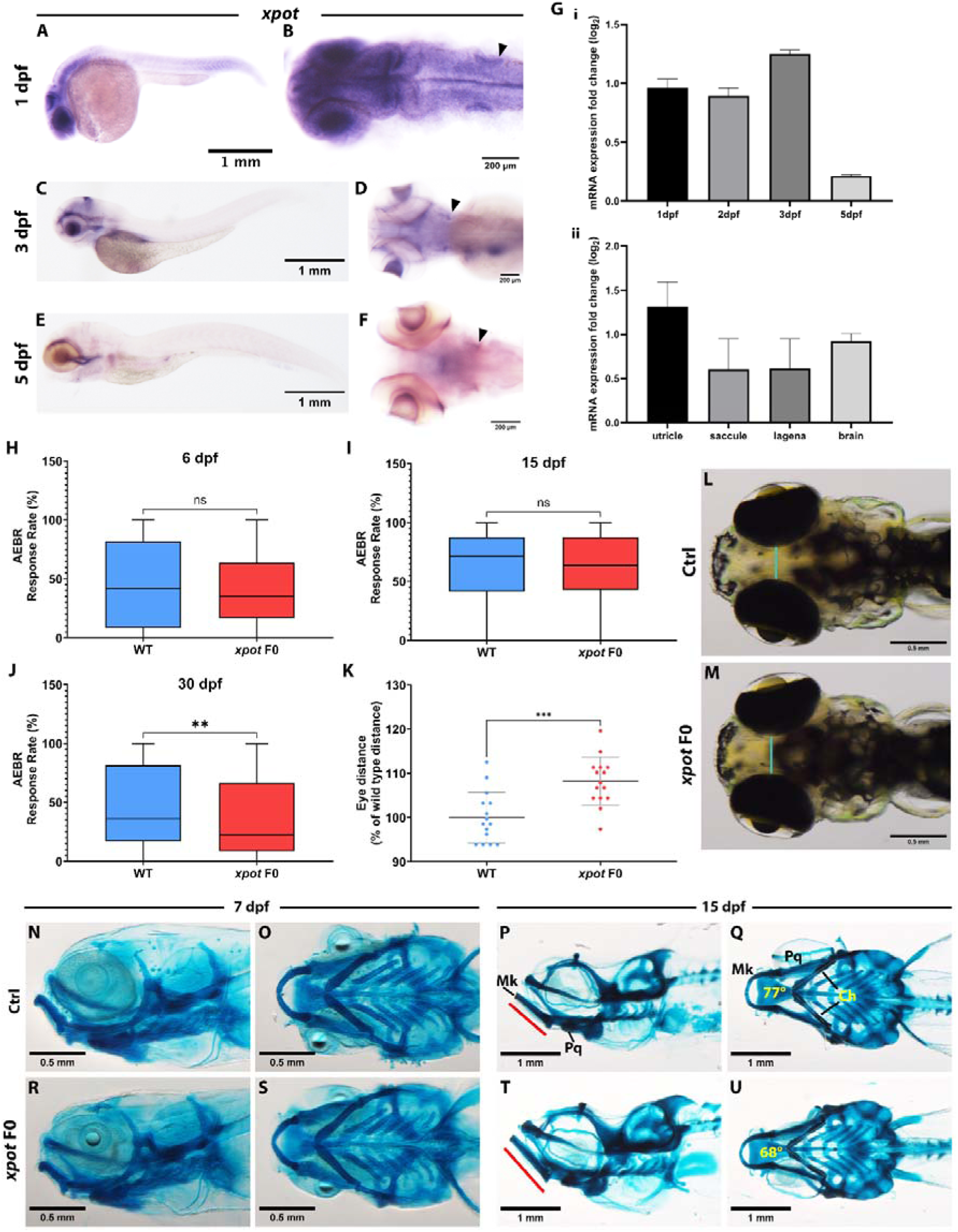
Zebrafish *xpot* knockout recapitulates key features of the human *XPOT* deficiency phenotype. **(A-F)** Whole-mount *in situ* hybridization showing *xpot* expression during zebrafish development. **(A, B)** At 1 dpf, *xpot* is broadly expressed throughout the embryo with a prominent signal in the head region, including the otic vesicle (arrowhead in B). **(C, D)** At 3 dpf, expression persists with enrichment in the developing inner ear (arrowhead in D). **(E, F)** At 5 dpf, *xpot* expression continues in cranial structures, including the otic region (arrowhead in F). Scale bars: 1 mm (A, C, E) and 200 µm (B, D, F). **(G)** Quantitative RT-PCR analysis of *xpot* expression. **(G-i)** Temporal expression profile showing *xpot* mRNA levels across developmental stages (1-5 dpf), with peak expression at 3 dpf. **(G-ii)** Tissue-specific expression in adult zebrafish, revealing the highest *xpot* levels in the utricle compared to the saccule, lagena, and brain. Data are presented as log_2_ fold change relative to the reference gene, *gapdh*. Error bars indicate SD. **(H-J)** Acoustic evoked behavioral response (AEBR) assay demonstrating progressive hearing impairment in *xpot* F0 animals. Response rates (percentage of responses to 12 acoustic stimuli) were measured at **(H)** 6 dpf, **(I)** 15 dpf, and **(J)** 30 dpf. No significant differences were observed at 6 and 15 dpf (ns), whereas *xpot* F0 larvae showed significantly reduced AEBR response rates at 30 dpf compared to wild-type (WT) controls. Data are presented as median with interquartile range; each dot represents an individual larva. **(K-M)** Hypertelorism in *xpot* F0 zebrafish. **(K)** Quantification of interocular distance normalized to wild-type mean, demonstrating significantly increased eye-to-eye distance in *xpot* F0 larvae compared to WT controls. Each dot represents an individual larva; horizontal lines indicate mean ± SD. **(L, M)** Representative dorsal view images of **(L)** control and **(M)** *xpot* F0 larvae illustrating increased interocular distance (cyan lines). Scale bars: 0.5 mm. **(N-U)** Alcian blue cartilage staining revealing craniofacial abnormalities in *xpot* F0 zebrafish. **(N-Q)** Control larvae at 5 dpf **(N, O)** and 10 dpf **(P, Q)** showing normal craniofacial cartilage development in ventral (N, P) and lateral (O, Q) views. **(R-U)** *xpot* F0 larvae at 5 dpf **(R, S)** and 10 dpf **(T, U)** displaying altered craniofacial cartilage morphology in ventral (R, T) and lateral (S, U) views. Scale bars: 0.5 mm (5 dpf) and 1 mm (10 dpf).

We then used CRISPR/Cas9 mutagenesis to induce indel mutations in *xpot*, and examined the phenotypic consequences in the founder generations (F0 knockouts, F0 KOs). The head and eye size of *xpot* F0 KOs were indistinguishable from Cas9-injected control animals by 5 dpf. Hearing was assessed at multiple developmental stages using the acoustic evoked behavioral response (AEBR) assay. At 6 dpf and 15 dph, the response rates of *xpot* F0 crispants were indistinguishable from wild-type controls (Figure 4H,I). However, by 30 dpf, *xpot* F0 larvae exhibited significantly reduced AEBR responses compared to controls (Figure 4J).

To assess craniofacial development in zebrafish, we quantified interocular distance as a measure of midfacial width. Strikingly, *xpot* F0 zebrafish displayed significantly increased eye-to-eye distance compared to wild-type siblings (Figure 4K). Representative dorsal images clearly illustrate the widened eye spacing in mutants compared to controls (Figure 4L, M). To further characterize the basis of craniofacial abnormalities, we performed Alcian blue cartilage staining. At 7 dpf, *xpot* F0 larvae showed subtle changes in craniofacial cartilage development compared to controls (Figure 4N-O and R-S). These differences became more pronounced by 15 dpf, with mutants displaying altered cartilage morphology, particularly a smaller angle between ceratohyal cartilage (ch) (Figure 4P-Q and T-U), and increased length of Meckel’s cartilage. Taken together, loss of Xpot function in zebrafish results in progressive hearing impairment and craniofacial abnormalities that closely parallel the clinical phenotype of patients with biallelic *XPOT* variants.

## Discussion

Herein we describe a previously unrecognized syndromic disorder caused by biallelic pathogenic variants in the tRNA exporter gene *XPOT*, implicating impaired nucleocytoplasmic transport of mature tRNAs as a disease mechanism in humans. The identified loss-of-function variants are predicted to result in nonsense-mediated decay, consistent with the complete absence of XPOT protein observed in patient-derived cells. Affected individuals share a remarkably uniform clinical presentation that includes recurrent infections, severe sensorineural hearing loss, and neurodevelopmental delay. A literature search identified five additional patients from a consanguineous family reported in the supplementary data of Hu et al. ^16^. According to the authors, these individuals exhibited mild psychomotor delay, moderate hearing loss, and speech impairment, and all carried the homozygous missense variant c.1850T>G p.(Met617Arg) in *XPOT*. This variant is located in the Armadillo-type fold and Exportin-t domain and affects an amino acid residue with a high degree of evolutionary conservation (phyloP: 7.59 [-19.0, 11.0]). It is predicted to be damaging by multiple tools (REVEL, CADD, PolyPhen2, SIFT), and the *XPOT* missense constraint Z-score is 3.29, which further supports increased intolerance to amino acid substitutions.

While disorders of tRNA biology have been linked to pathogenic variants in tRNA modifying enzymes and ARSs^17, 18^, a primary defect in tRNA nuclear export has, to our knowledge, not previously been linked to Mendelian disorders. Our genetic and functional data place the tRNA exporter at the center of a convergent pathomechanism that attenuates protein synthesis and perturbs cellular stress responses.

Biosynthesis of TNFα is significantly impaired in patients due to a reduced protein output per unit mRNA as reflected by a shallower protein–mRNA regression slope. This is most likely attributable to reduced translational efficiency, potentially reflecting limited availability of cytoplasmic tRNAs. Simultaneously, RNA-seq did not reveal a coordinated reduction of ARS transcripts, arguing against am impairment of the tRNA-charging machinery. Interestingly, the clinical features of patients with ARS deficiencies, which include central nervous system dysfunction, impaired hearing and/or vision or failure to thrive,^19^ overlap with features seen in *XPOT* deficiency. Liver dysfunction, pulmonary fibrosis, and neuropathy have also been reported in ARS deficiencies, some of them leading to early death. The phenotype in our *XPOT*-deficient patients is less severe, possibly due to partial compensation by alternative tRNA export mechanisms.

Inhibition of tRNA export through the introduction of antibodies against XPOT in *Xenopus* oocytes reduced cytoplasmic tRNA levels by approximately 80%^8, 20^, supporting that XPOT is the major, if not the sole, exportin required for active tRNA nuclear export in vertebrates. In contrast, XPOT is non-essential in several simple model organisms, including *Saccharomyces cerevisiae*, *Schizosaccharomyces pombe*, and *Arabidopsis thaliana*^21^.

Recent studies hypothesized that XPO1 (CRM1), the major nuclear export receptor for RNAs and proteins, and XPO5, which is primarily known for exporting miRNAs, are exportins capable of binding specific tRNAs and mediating their nuclear export^22,23^. Although others have suggested their contribution is minor^24, 25^. Additional candidates include NXF1 and NXT1 that could function in tRNA export^26^ and binding^27^. The marked reduction of transcription and translation of TNF-alpha in our patient-derived cells indicates that alternative export receptors cannot fully compensate for the loss of XPOT. Suprisingly, the reduction of proliferation in patient and control cells was not aggravated by an additional XPO1 inhibition. Therefore, further studies are required to determine which exporter compensates the function of XPOT in our patients. Passive diffusion may be an additional mechanism under conditions of elevated nuclear tRNA accumulation. Notably, a recent study identified heterozygous pathogenic variants in *XPO1* as a cause of monogenic neurodevelopmental disorders with autosomal-dominant inheritance. Importantly, none of the heterozygous carriers of the *XPOT* variants in our families have been diagnosed with a neurodevelopmental disorder ^28^.

The cellular consequences of impaired tRNA export appeared to be context dependent. It is plausible that tRNA availability is a temporally-sensitive process and that the principal consequence of XPOT deficiency is impaired ability to sustain rapid and efficient protein synthesis under conditions of increased demand, thereby contributing to weakened immune defence and the progressive hearing loss observed in patients with clear loss-of-function variants (P1-P6, P8). We hypothesis that limited tRNA availability attenuates protein turnover in the highly burdened, non-regenerative hair cells and spiral ganglion neurons, thereby leading to progressive structural, functional and/or degenerative changes resulting in hearing loss. Moreover, the milder phenotype observed in P7 and in the patients reported in the Supplementary data of Hu et al.^16^ may be explained by the presence of variants that still allow for residual *XPOT* function (c.270+1G>A and c.1850T>G). Although stereocilia exhibit exceptionally high protein turnover due to the continuous renewal of actin bundles and associated myosin motors, they lack ribosomes and therefore do not support local protein synthesis. All stereociliary proteins are synthesized in the hair cell cytoplasm and subsequently transported into the stereocilia. The pronounced enrichment of XPOT in stereocilia thus suggests a non-canonical, translation-independent function, that will need to be addressed in future studies.

Our zebrafish model provides critical *in vivo* validation of the pathogenicity of XPOT loss-of-function. The *xpot* F0 mutant zebrafish recapitulated two key features of the human phenotype, i.e. progressive hearing impairment and craniofacial dysmorphism. The temporal pattern of hearing loss in zebrafish, with normal responses at 6 and 15 dpf but a decline by 30 dpf, reflects the clinical observation that at least P2 and P3 were reported to clearly react to sounds in early infancy but lost hearing in their first years of life. This progressive decline supports our hypothesis that XPOT deficiency leads to cumulative translational deficits in metabolically demanding cells such as hair cells and spiral ganglion neurons. The hypertelorism observed in *xpot* F0 zebrafish provides a developmental correlation to the midface hypoplasia and broad nasal bridge present in 5/7 patients.

Our study has some limitations. This is a retrospective multicentre study, and not all clinical parameters and diagnostic findings were available. In particular, it would have been useful to perform systematic and standardised immunological investigations on all patients. Systematic virological, bacteriological and mycological examinations of the lower respiratory tract to identify dominant pathogens would also be beneficial, particularly with regard to possible infection prevention approaches. A prospective observational cohort study would be highly desirable in this regard.

Furthermore, it will be important to understand the immunological mechanisms involved. It also remains unclear which specific immune pathways are particularly affected by the tRNA exporter defect. The analyses of the patients described here did not reveal any defects in B- or T-cells. In addition, immunoglobulin levels and antibodies, when tested, were largely within normal ranges.

Similarly, the zebrafish model provides valuable functional validation, but several limitations exist. First, phenotyping was performed in F0 knockout animals, which exhibit mosaic gene disruption. Although our data demonstrate substantial similarity between F0 knockouts and stable genetic mutants^29^, the variability observed may not capture subtle phenotypes, and a stable knockout is necessary to fully characterize these phenotypes and enable further mechanistic studies. Second, zebrafish lack a cochlea and instead possess inner ear structures highly similar to those of humans^30^, while the AEBR assay robustly measures auditory function, it may not capture the full complexity of sensorineural hearing loss observed in patients. Additionally, key aspects of the human phenotype, including recurrent respiratory infections, bronchiectasis, and immune dysregulation, cannot be readily modeled in zebrafish due to fundamental anatomical and immunological differences. Despite these limitations, the concordance between zebrafish and human phenotypes supports a strong genotype-phenotype relationship and provides a tractable model system for further mechanistic studies.

In summary, we define *XPOT* deficiency as a monogenic disorder caused by potentially defective tRNA export, thereby linking disruption of a core nuclear transport pathway to human disease (Figure 5). Understanding how XPOT loss perturbs translation in distinct cell types will not only clarify disease mechanisms but may also highlight new targets for restoring proteostatic balance in affected tissues.

**Figure 5:**
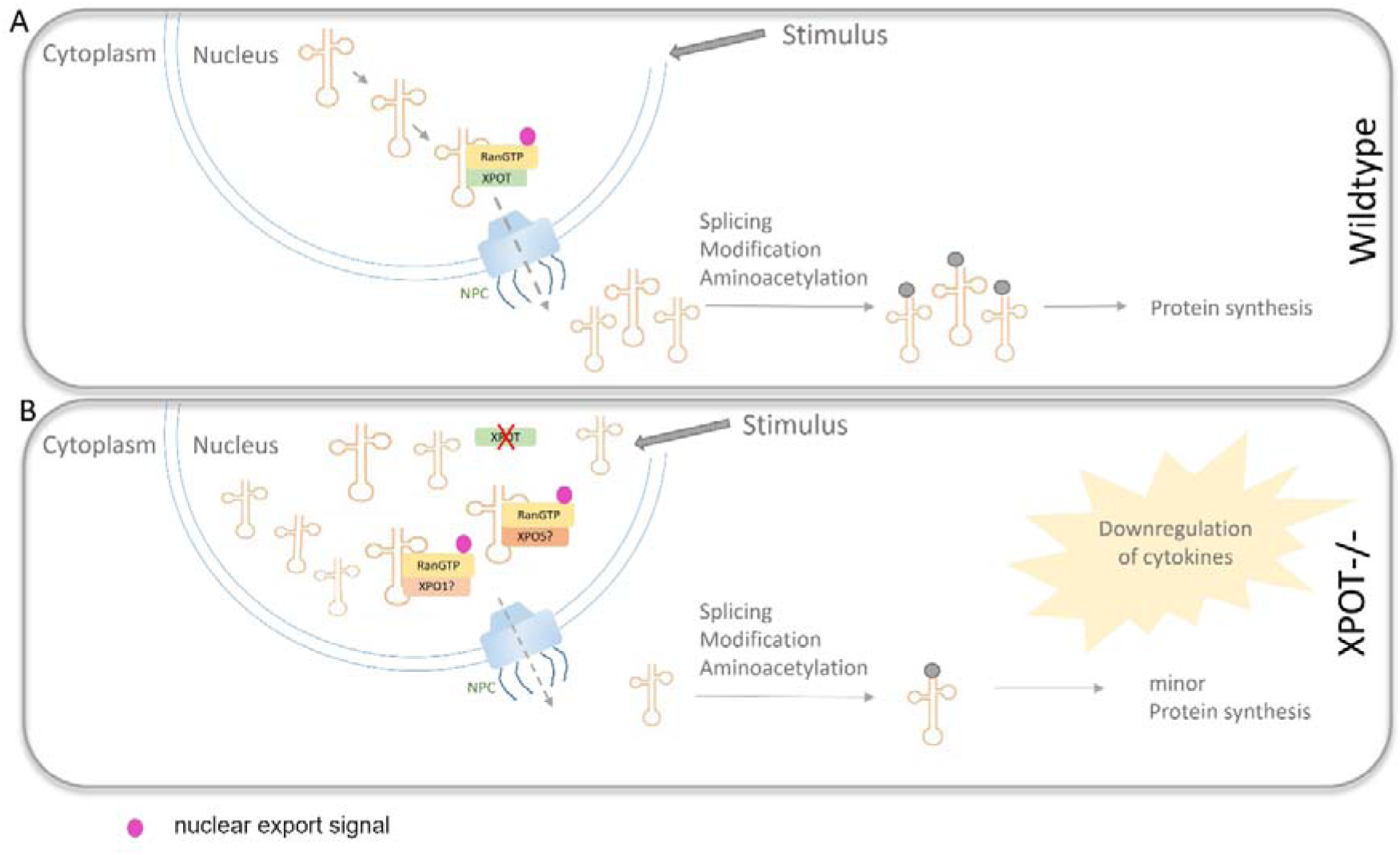
A: Schematic representation of the pathogenic mechanism of XPOT. The tRNA genes are located in the nucleus and are transcribed by RNA polymerase III. The resulting pre-tRNA is processed within the nucleus to form mature tRNA (not shown). Subsequently, export from the nucleus occurs via the tRNA export receptor XPOT after binding a nuclear export signal through a RanGTP-dependent mechanism across the nuclear pore complex (NPC). After export, additional modifications may occur before the tRNA is charged with its corresponding amino acid by an aminoacyl-tRNA synthetase and binds to the ribosomal complex for protein synthesis. **B:** In XPOT deficient (XPOT-/-) cells, mature tRNAs cannot exit the nucleus. This leads to an accumulation of tRNAs within the nucleus and a corresponding depletion in the cytoplasm. As a result, translation cannot proceed efficiently, causing a reduction in protein synthesis. Nevertheless, an alternative export pathway appears to maintain a minimal level of protein biosynthesis necessary for basic cellular function. Export receptors such as XPO1 (CRM1) or XPO5 could mediate a low level of tRNA export; however, a compensation seems not efficient enough to fully replace XPOT.

## Methods

### Study information

Written informed consent was obtained from the legal guardians of the patients at the respective institutions. The study was approved by the Ethics Committee (AZ_8657_BO_K_2019) at Hannover Medical School. All procedures involving experimental animals were conducted in accordance with the institutional policies, including the University Medical Center Göttingen Animal Welfare Office (ref. T22-42, mouse) and NIH guidelines (IACUC; protocol #24-28, zebrafish).

### Genetic and Functional Analyses

Details of the genetic and functional analyses, including the mouse and zebrafish experiments, can be requested directly from the corresponding author.

## Data Availability

All data produced in the present study are available upon reasonable request to the authors

## Acknowledgments

We would like to thank the families for their participation. We thank Dr. Reza Azizimalamiri, Dr. Gholamreza Shariati, Dr. Hamid Galehdari, Niloofar Chamanrou, Dr. Tahere Seifi, and Jawaher Zeighami for clinical and genetic data collection. We thank Michaela Losch, Bernd Haermeyer and Marlies Eilers for their excellent technical assistance.

This work was supported by the German Research Foundation (DFG) Heisenberg program (STR1027/5-2 to N.S. and VO2138/8-1 grant 543719215 to B.V.), and the DFG Collaborative Research Center 1690 (Project A01 to N.S. and A03 to B.V.). This work was done with the support of the Center for Rare Hearing Disorders at the Center of Rare Diseases Göttingen (ZSEG). Zebrafish work is supported by the Oklahoma Medical Research Foundation, Oklahoma City, USA.

## Author contributions

SvH, IN, NDD, BA, GS, SLK and GKV Contributed to the study conception and design.

Experiments and data collection were performed by SvH, ID, VS, JR, KH, CP, SS, SSe, SJL, AE

Patient recruitment, sample collection, and clinical assessment were conducted by AW, TR, NSch, CP, ALB, SG, KW, RA, AS, SE, RA & AS

WES, WGS, bioinformatics and/or data analysis were performed by SvH, MZ, SK, SB, SK

Supervision was provided by BV, NDD, NSt, BA, and GKV.

The first draft of the manuscript was written by SvH, IN, BV, and GKV.

## Declaration of interests

The authors have no conflicts of interest relevant to this article to disclose.

## Notes

### Competing Interest Statement

The authors have declared no competing interest.

### Author Declarations

The study was approved by the Ethics Committee (AZ_8657_BO_K_2019) at Hannover Medical School.

