## Supplementary Appendix for "*XPOT* Deficiency causes a human disorder through impaired tRNA nuclear export"

| **Reagent type, species or resource** | **Designation** | **Source of reference** | **Identifiers** | **Additional information** |
| --- | --- | --- | --- | --- |
| **Immunohistochemical analyses of XPOT in the mouse cochlea** | | | | |
| Primary Antibody | Anti-XPOT (rabbit polyclonal) | Thermo Fisher Scientific/ Invitrogen | Cat# PA5-88317  RRID AB_2804824 | Dilution for IHC: 1:200 |
| Primary Antibody | Anti-XPOT (rabbit polyclonal) | Thermo Fisher Scientific/ Invitrogen | Cat# PA5-65492  RRID AB_2662125 | Dilution for IHC: 1:200 |
| Primary Antibody | Anti-Parvalbumin (mouse monoclonal) | Synaptic Systems | Cat# 195011  Lot# 1-6  RRID AB_2619882 | Dilution for IHC: 1:200 |
| Chemical compound | Phalloidin Alexa Fluor 633 | Thermo Fisher Scientific/ Invitrogen | Cat# A22284  Lot# 2604076 | Dilution for IHC:  1:500 |
| Chemical compound | Hoechst 34580 | Thermo Fisher Scientific/ Invitrogen | Cat# H21486  Lot# 2604340 | Dilution for IHC: 1:1000 |
| Secondary Antibody | Anti-rabbit Alexa Fluor 488 (donkey polyclonal) | Thermo Fisher Scientific/ Invitrogen | Cat# A-21206  Lot# YH374677  RRID AB_2535792 | Dilution for IHC: 1:200 |
| Secondary Antibody | Anti-mouse Alexa Fluor 568 (goat polyclonal | Thermo Fisher Scientific/ Invitrogen | Cat# A-11004  Lot# 2633533  RRID AB_2534072 | Dilution for IHC: 1:200 |
| Sample preparation reagent | Epredia™ Cryomatrix™ embedding resin | Thermo Fisher Scientific, Fisher Scientific | Cat# 67-690-06 |  |
| **Western blot analysis of XPOT in fibroblasts/LCLs** | | | | |
| Primary Antibody | Anti-XPOT (rabbit polyclonal, IgG) | Thermo Fisher Scientific/ Invitrogen | Cat# PA5-66095  RRID AB_2662126 | Dilution for WB: 1:500 |
| Secondary Antibody | Anti-rabbit IRDye® 800CW (donkey polyclonal, IgG) | IRDye® 800CW | Li-Cor, 926-32213, RRID: AB_621848 | Dilution for WB: 1:15000 |
| **Immunofluorescence analysis of XPOT in fibroblasts** | | | | |
| Primary Antibody | Anti-XPOT (rabbit polyclonal, IgG) | Thermo Fisher Scientific/ Invitrogen | Cat# PA5-66095, RRID:AB 2662126 | Dilution for IF: 1:300 |
| Primary Antibody | Anti-γCYA (mouse monoclonal, IgG2b) | Bio-Rad | Cat# clone 2A3, RRID:AB_2571583 | Dilution for IF: 1:500 |
| Secondary Antibody | Anti-rabbit Alexa Fluor 555 (donkey polyclonal) | Thermo Fisher Scientific/ Invitrogen | Cat# A-31572, RRID:AB_ 162543 | Dilution for IF: 1:500 |
| Secondary Antibody | Anti-mouse Alexa Fluor 488, IgG, Fcγ Subclass 2b Specific (goat polyclonal) | Jackson Immunoresearch | 115-175-207, RRID: AB_2338717 | Dilution for IF: 1:500 |

**Supplement Table S1: Antibodies used for Immunohistochemical analyses of XPOT in the mouse cochlea, Western blot analysis and Immunofluorescence analysis**

**Supplement Table S2: Primers used in the study**

| **Primer name** | **Primer sequence (5´-3´)** | **Purpose** |
| --- | --- | --- |
| hu_XPOT Ex 5 XhoI F | aattctcgagCATGGGCACTTGTGTTCAGA | Amplification of gDNA and cloning |
| hu_XPOT Ex 5 BamHI R | attggatccAGAGCCAAACAGAAAGCAGC | Amplification of gDNA and cloning |
| XPOT_c.270+1G-A_mut_F | GCAAGCTCAGaTAAAATCATAATTTC | Site-directed mutagenesis |
| XPOT_c.270+1G-A_mut_R | AGCCATGATATGAGCGTC | Site-directed mutagenesis |
| SD6 F | TCTGAGTCACCTGGACAACC | Colony PCR and RT-PCR |
| SA2 R | ATCTCAGTGGTATTTGTGAGC | RT-PCR |
| Xpot_T1 | GCGCTGAAGCCCTCGCTAAA | sgRNA for CRISPR editing |
| Xpot_T2 | CGACATCCTACACAGCCCCG | sgRNA for CRISPR editing |
| Xpot_T3 | AATGAGATTGAGGTCGATCC | sgRNA for CRISPR editing |
| T3-xpot-F1 | GaattgaattaaccctcactaaagggGCTTACCTGTTCTCACGCTT | WISH |
| T7-xpot-R1 | gaattgtaatacgactcactatagggATCCGGAAATCCCACCAAAC | WISH |
| xpot_qpcr_F | GACACGCCACTGTTCGATCT | qPCR |
| xpot_qpcr_R | GAAATATGCCAGCGCCCTCT | qPCR |
| Xpot_T1F | TCATACATTGCATCCTAATAGAGCT | Genotyping |
| Xpot_T1R | CTCCAAGACTTGAAAGCAGAAGA | Genotyping |
| Xpot_T2F | GTGTGTTTGTGCAGCTGATGAA | Genotyping |
| Xpot_T2R | ATCGTGACGTATGGTTTACTGT | Genotyping |
| Xpot_T3F | TCTGTTGTTCACTAGGAGACCC | Genotyping |
| Xpot_T3R | AGCTGAATACTAGTAACTTGAAGGA | Genotyping |

**Supplement Table S3:** Table of the genetic variants in the patients, including sex, consanguinity, and ethnicity; * = patients from the paper by Hu et al.

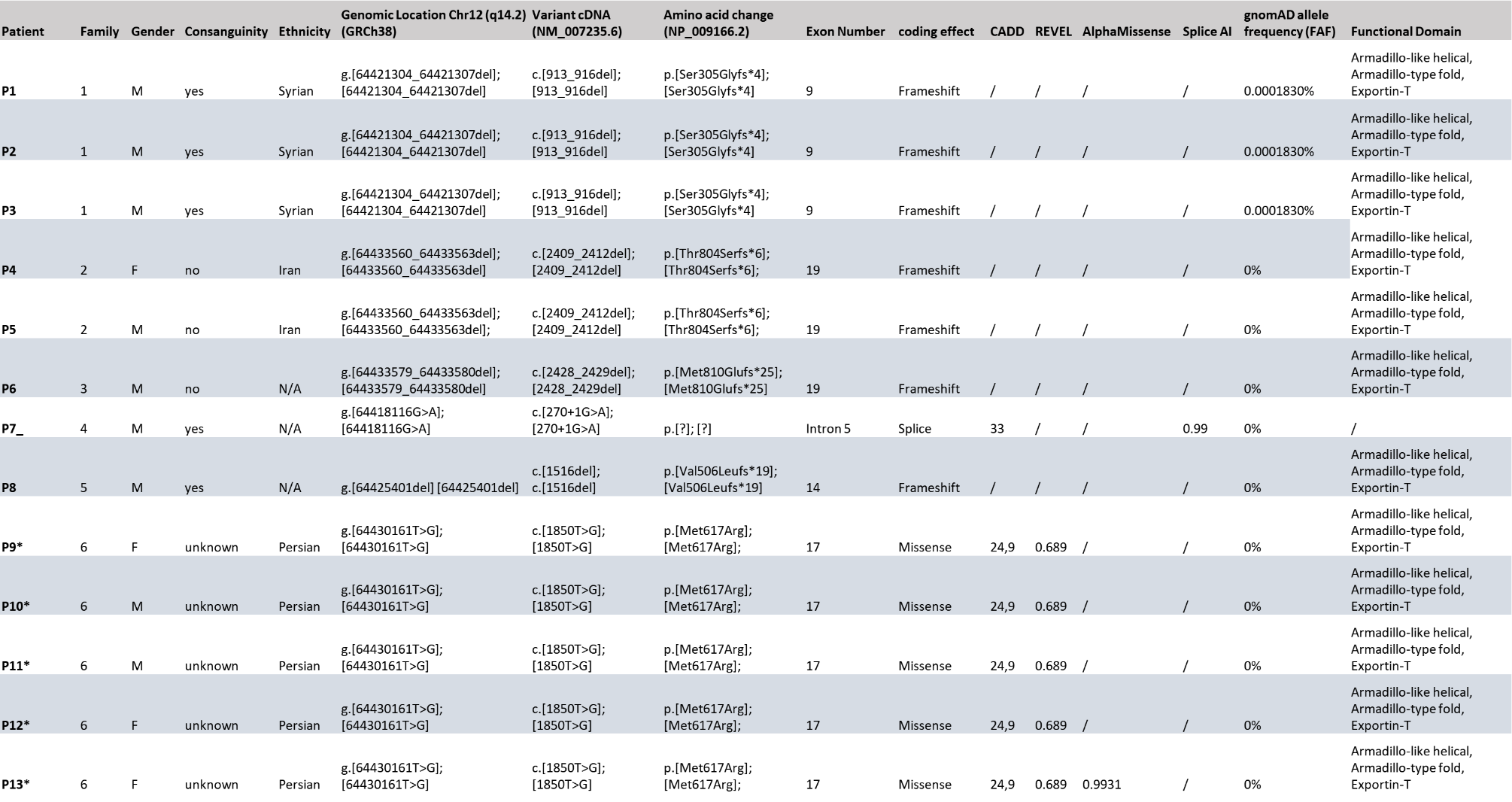

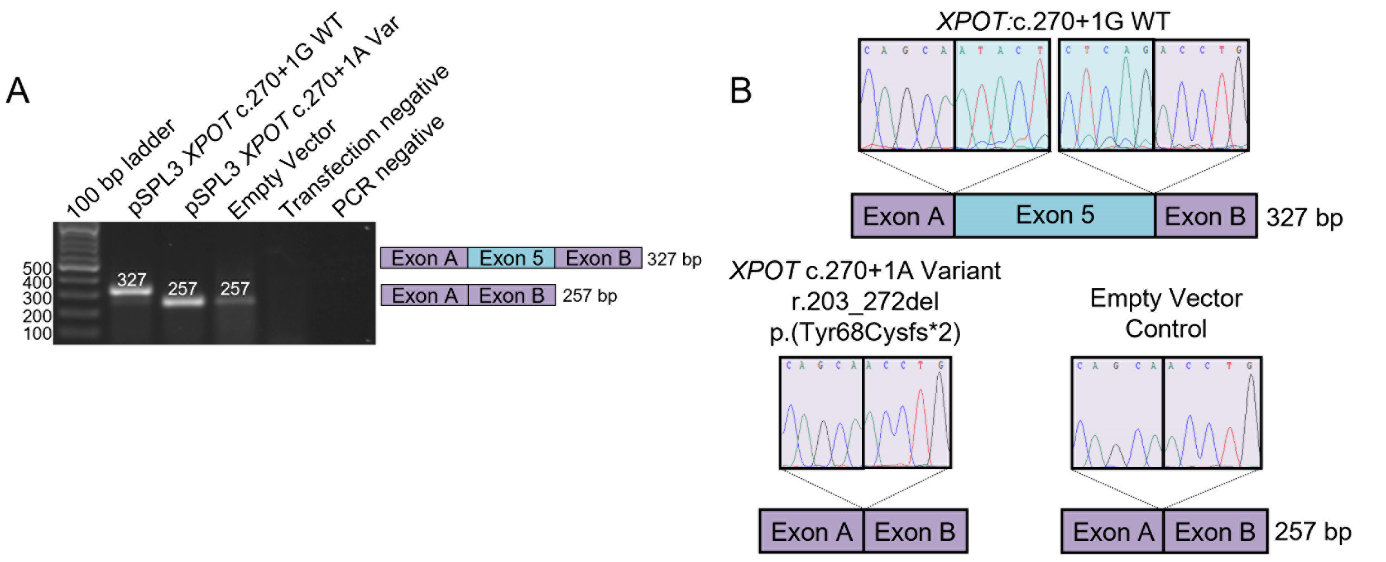

**Supplement Figure S1:** Splicing schematic for the *XPOT* c.270+1G>A variant. **A**: Gel electrophoresis of the RT-PCR from the wild-type control (c.270+1G), the c.270+1A variant, as well as the empty pSPL3 vector, transfection negative, and PCR-negative controls. The graphic to the right shows the wild-type result, corresponding to the 327 bp band. The variant shows evidence of exon skipping (257 bp), identical to the empty vector control. **B**: Sanger sequencing confirmation of the RT-PCR amplicons. The upper panel shows the wild-type sequence of exon-exon junctions. The lower panels show Sanger sequencing of the exon 5–skipped transcript resulting from the c.270+1G>A variant, which leads to a frameshift (r.203_272del, p.(Tyr68Cysfs*2)), and the empty vector control (left and right, respectively).

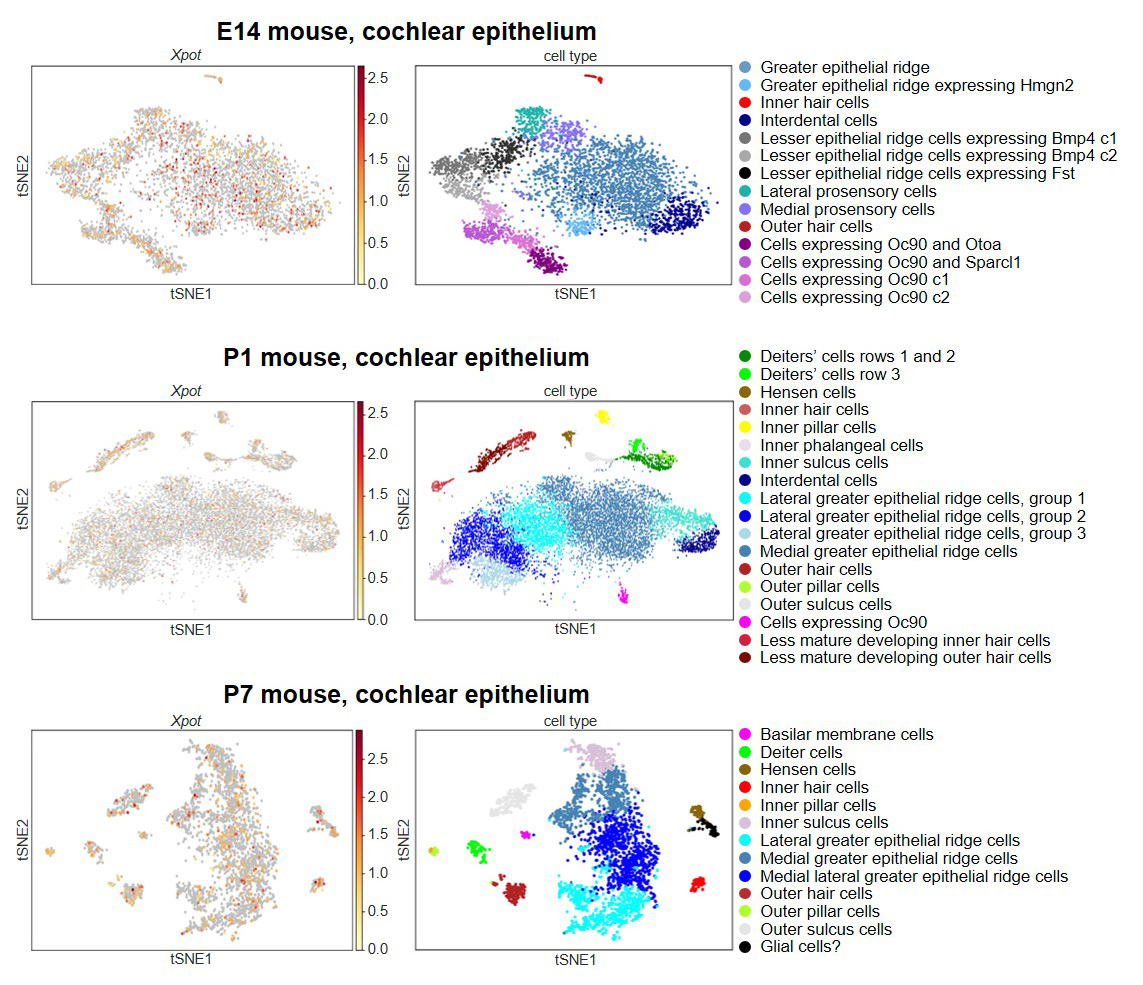

**Supplement Figure S2:** Expression of XPOT in the mouse cochlea through RNA-seq data. **A** Visualization of *Xpot* expression in the embryonic (E14, upper panel) and postnatal (P1 and P7, middle and lower panel, respectively) mouse cochlear epithelium using single cell RNA-seq data. Data were visualized using the gene Expression Analysis Resource (gEAR) portal.  Inner ear cochlear expression data are available from gEAR https://umgear.org/expression.html

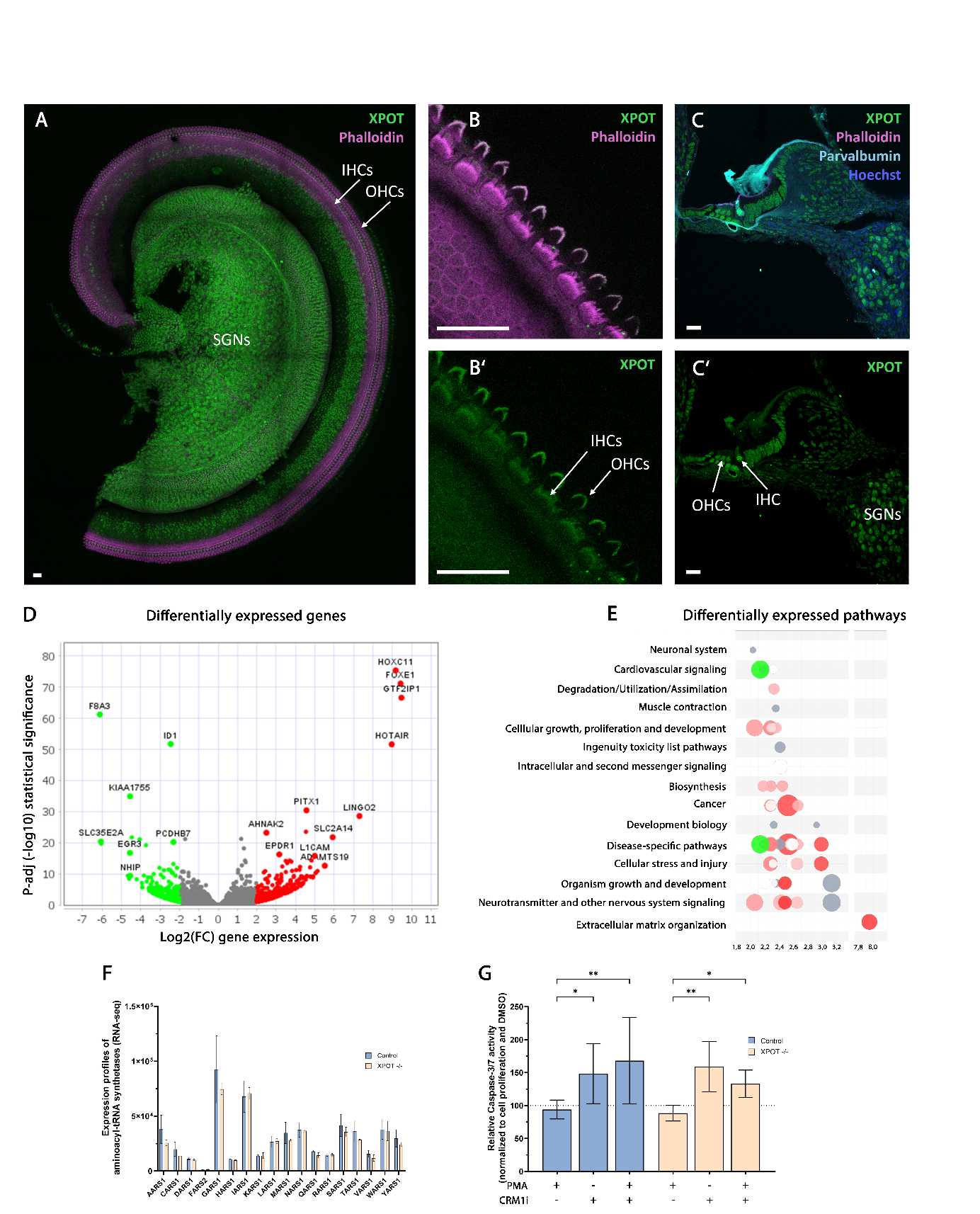

**Supplement Figure S3**: **A**: Maximal intensity projections of confocal images of immunostaining of a whole mount preparation of an organ of Corti of a wildtype mouse (p5) showing ubiquitous cytoplasmic and nuclear expression of XPOT (green), including spiral ganglion neurons (SGNs) and inner and outer hair cells (IHCs, OHCs). **B**: Higher magnification of IHC and OHC stereocilia. Fluorophore-coupled phalloidin was used to counter-stain actin in stereocilia and supporting cells (cyan). In B, the stereocilia of the second and third row of outer hair cells are not in the plane of focus. **C**: Cryosection of an organ of Corti of a wildtype mouse (p28) with ubiquitous cytosolic XPOT labelling (green), including IHCs and type I SGNs (co-labeled parvalbumin). Nuclei are stained with Hoechst. All scale bars in A-C are 20 µm. All XPOT stainings in this figure were performed using antibody #PA5-65492 from Invitrogen, whereas #PA5-88317 was used for the images in main figure 3. **D** Volcano plot displaying genes significantly upregulated (green) and downregulated (red) in two mutant fibroblasts compared to four WTs under steady-state condition. **E**: Bubble plot illustrating gene ontology (GO) enrichment analyses of downregulated genes in *XPOT* mutant fibroblasts compared to WTs. Only pathways with a –log(p-value) greater than 2.0 were included. Entities were sorted by –log(p-value). In the chart, the vertical and horizontal axes represent –log(p-value). Bubble size corresponds to the number of genes overlapping with the pathway. Bubble colour reflects the z-score, indicating negative (blue), zero (white), or positive activity patterns (red); pathways without an identifiable activity pattern are shown in grey. The bubble chart is based on Fisher’s exact test p-values. **F**: Expression of aminoacyl-tRNA synthetases. Data are the mean of four control and two patient fibroblast cell lines. Error bars indicate SD. **G**: Quantification of fibroblast caspase 3/7 activity after PMA, Selinexor or combined PMA and Selinexor treatment as a percentage relative to DMSO-treated controls. Data are the mean of four control cell lines and two patients cell lines (P2, P3), each from three independent assays. Error bars indicate SD; *, P < 0.05; **, P < 0.01 (Kruskal-Wallis test).

**Detailed description of audiological development in patients with *XPOT* deficiency:**

*Patient 1 (P1)*

Hearing impairment was suspected in early childhood. The patient was later briefly fitted with bilateral hearing aids. Between 5 and 10 years of age, Auditory Brainstem Response (ABR) testing showed no reproducible responses to click and chirp stimuli. He received high-power hearing aids and barely responded to sounds two years later. Transient evoked otoacoustic emissions (TEOAE) were absent. Temporal bone CT revealed normal anatomical configuration of the middle and inner ear structures bilaterally. Cranial MRI showed no evidence of pathology of the vestibulocochlear nerves. Cochlear implantation of the right ear was performed. Together with intensive auditory rehabilitation, this restored his open field thresholds to 20-40dB and he achieved 70%-word perception at 65dB (Mainzer I). With the hearing aid in the left ear, his thresholds ranged between 40 and 60dB and word perception at 80dB was 50%. Prompted by the good success of cochlear implantation, his left ear was implanted. Afterwards, P1 was able to reliably distinguish everyday environmental sounds and perceive spoken language in his surroundings. Open field thresholds with either CI were around 30dB. Between 10 and 15 years of age, he had 60% correct word perception in a closed set test (Mainzer II) with either CI. His expressive language remained limited to a few intelligible words.

In summary, P1 has bilateral severe prelingual cochlear sensorineural hearing loss. Hearing aids did not suffice for speech acquisition, whereas cochlear implantation significantly improved sound detection skills and enabled limited speech development, mainly supporting his use of speech-accompanied sign language.

*Patient 2 (P2)*

P2 initially clearly responded to sounds but developed obvious hearing impairment. Tympanostomy tubes were inserted twice because of chronic middle ear effusion. Sensorineural hearing loss was diagnosed based on ABRs, with only residual hearing in the right and severe hearing loss in the left ear. Bilateral hearing aids were prescribed and were accepted well. Free-field audiometry at routine frequencies between 0.25 and 8 kHz showed pancochlear responses between 80 and 90 dB without and between 60 and 70dB with hearing aids. TEOAE were absent. ABRs where thresholds were at least 90dB on the left side, and mostly missing on the right side. Tympanograms showed variable results and stapedial reflexes were absent. In CT and MRI scans, the temporal bones, cochleae, semicircular canals, ossicles and the eight cranial nerve appeared normal. He received a cochlear implant for the right ear between 5 and 10 years. He accepted the speech processor very well and obtained open field thresholds of 35-40dB. However, he developed little speech perception. Cochlear implantation followed on the left ear. In summary, P2 suffered from prelingual cochlear sensorineural hearing loss, which is profound in the right and severe in the left ear. In addition, he had recurrent middle ear effusion. Hearing aids were beneficial but not sufficient for speech development. Cochlear implantation was successful, although the other syndromic features and the relatively late diagnosis and rehabilitation limit the development of communication skills.

*Patient 3 (P3)*

- ABR of P3 showed bilateral, pancochlear moderate to severe hearing loss in early childhood. He was fitted with hearing aids and afterwards free-field responses were at 60-70 dB between 0.5-4 kHz and improved to 30-40 dB over the same frequencies with hearing aids. Nevertheless, hearing loss progressed. He did not react to sounds anymore and ABRs disappeared. He received bilateral cochlear implants and reacts to medium intensity sounds but does not seem to understand speech. He uses limited sign language and is not yet able to participate in subjective audiometry tests.

In summary, P3 had mild/moderate sensorineural hearing loss at birth which progressed to severe hearing loss and later in childhood to deafness. He never had clear benefit from hearing aids. With cochlear implants, he perceives sounds. Speech rehabilitation is limited by cognitive and motor dysfunction and by recurrent hospitalizations due to lung disease.

*Patient 4 (P4)*

- An audiogram between 30 and 35 years of age shows profound deafness in the right ear. In the left ear, bone conduction thresholds are 60-65dB. Air conduction thresholds are 70-90dB between 250 Hz and 2kHz and >110dB at 4 and 6kHz. The audiogram shows a slight notch at 1kHz. The patient relies on sign language.

*Patient 5 (P5)*

- An audiogram between 30 and 35 years of age shows bilateral profound deafness. The patient relies on sign language.

*Patient 8 (P8)*

- The patient was reported to have early-onset hearing loss with thresholds of 80 dB in the left and 60dB in the right ear. He uses hearing aids.
